# Breast Cancer Trends in Chile: Incidence and Mortality Rates (2007–2018)

**DOI:** 10.1101/2022.11.04.22281953

**Authors:** Benjamín Madariaga, Susana Mondschein, Soledad Torres

## Abstract

**Introduction:** Breast cancer (BC) is one of the most common cancers in women worldwide and in Chile. Due to the lack of a Chilean national cancer registry, there is partial information on the status of breast cancer in the country.

**Aims:** To estimate breast cancer incidence and mortality rates by health care providers and region for Chilean women.

**Methods:** We used two public anonymized databases provided by the Department of Health Statistics and Information of the Ministry of Health: the national death and hospital discharges datasets. For calculations of incidence and mortality rates, we selected all patients whose primary diagnosis was breast cancer, according to the CDI-10 diagnostic code.

**Results:** We considered a cohort of 58,254 and 16,615 BC hospital discharges and deaths for the period 2007–2018. The new cases of BC increased by 43.6%, from 3,785 in 2007 to 5,435 in 2018. Total BC deaths increased by 33.6% from 1,158 to 1,547 during the same time period. Age–adjusted incidence rates were stable over time, with an average rate of 44.0 and a standard deviation of 2.2 during the study period. There were considerable differences in age–adjusted incidence rates among regions, with no clear geographical trend. Women affiliated to a private provider (ISAPRE) have an average age adjusted incidence rate of 60.6 compared to 38.8 for women affiliated with the public provider (FONASA). Age–adjusted mortality rates did not change significantly during the study period, with an average of 10.5 and a standard deviation of 0.4.

**Discussion:** This study shows important differences in incidence rates between private and publicly insured women, with no significant differences in mortality rates. This result suggests potential inequalities in breast cancer healthcare outcomes in Chilean women, that should be studied in greater depth. Additionally, differences in breast cancer incidence found in this study compared to incidences reported from other estimations reinforce the need of a national cancer registry that should lead to more accurate indicators regarding breast cancer magnitude in Chile.

## 1 Introduction

Breast cancer (BC) is one of the most common cancers in women worldwide, with 2.26 million new cases and 685 thousand deaths globally in 2020 and 7.8 million women alive with breast cancer diagnosed in the past 5 years [1]. In 2020, age-standardized world breast cancer incidence and mortality rates were 47.8/100,000 and 13.6/100,000 women, respectively [2], while the figures reported for Chile by the Ministry of Health (Minsal) for 2018 were 44.1/100,000 and 11.3/100,000 women for incidence and mortality rates, respectively [3, 4].

Unfortunately, in Chile, there is no national cancer registry, and thus, incidence rates have been estimated based on projections on the number of breast cancer cases diagnosed from 1998 to 2012, with follow-ups until 2015, from four regional population-based registries (Antofagasta and Los Ríos regions and Concepción and Biobío provinces). The estimates for the rest of the country and from 2013 onward are based on statistical models, with assumptions such as a constant fatality rate across regions and over time, and in some instances, using neighboring country data [3, 5]. However, these assumptions might lead to unreliable estimates when ignoring significant differences in ethnicity, urbanization, socioeconomic composition and health coverage among regions [6–9] and advances in breast cancer treatment over time. Thus, for example, fatality rates might largely vary among regions due to differences in incidence as well as in mortality rates and over time due to a decrease in mortality rates.

On the other hand, mortality rate estimates are much more reliable because of the existence of a high-quality national death registry that includes a series of demographic variables along with the cause of death of every decedent in Chile. The death registry has been used in studies such as those by Icaza, Nuñez and Bugueño [10], where the authors exploit the potential of this registry by studying breast cancer mortality, including ecological analysis by sociodemographic variables.

The Chilean health care system consists of a hybrid of public and private insurance plans as well as providers. The three main insurance systems are the National Health Fund (FONASA) for 78% of the Chilean population, private health care insurers (ISAPREs) for 14% of the Chilean population, and the Military and Police Forces’ health system for 3% of the Chilean population. The remaining 5% of the population either has no health care insurer or their insurance status is unknown. Privately insured patients can solely access private providers, with a variety of coverage, depending on their health care plan and monthly payments. On the other hand, FONASA patients – paying, on average, a lower monthly price compared to those in the private system, might access public and private hospitals, depending on the health care plan within FONASA, with different amounts of copayments. Thus, the FONASA has four different segments of patients (A, B, C, and D), with A and D being those plans with the lowest- and highest-income patients, respectively. FONASA also provides coverage for attending private health centers to patients from the B, C and D segments [11].

The selection of the health care provider is determined mainly based on the individual’s economic income [12, 13]; as a consequence, profound inequalities in terms of quality of life and education, among others, have existed between users of the private and public affiliates. Health care indicators have been significantly better for those with private health insurance compared to those for publicly insured patients. For example, in 2017, 27.8% of FONASA affiliates declared having problems obtaining health care, while this percentage decreased to 12.6% for ISAPRE affiliates [6].

The goal of this paper is the estimation of breast cancer incidence and mortality rates, at a national and regional levels using an alternative methodology, based on a complete anonymized public database of national registry of hospital discharges, which includes information and diagnosis at the patient level, and the national death registry, with primary and secondary causes of death through a unique ID classification, compiled by the Department of Health Statistics and Information (DEIS) of the Ministry of Health. We also compute incidence and mortality by healthcare insurance system.

## 2 Methods and Materials

### Ethics statement

This work used publicly available data from the Chilean Ministry of Health through the Department of Health Statistics and Information. All data were protected, and personal information was anonymized; therefore, no consent from participants was needed.

### 2.1 Data

Two public anonymized databases were provided by DEIS. The first is the National Death Registry, which includes 2,549,800 deaths from January 1990 to December 2018. For each death entry in the registry, the patient’s ID (identifying code), date of birth, date of death, gender, town and region of residence, marital status, occupation and cause of death code according to the International Statistical Classification of Diseases and Related Health Problems (CDI-10) were available. There were 2,984 death registries without an ID, which corresponds to 0.1% of the complete database. The second database includes all discharges from public and private health care facilities in the country, which consists of 32,443,591 registries from January 2001 to December 2020. Each registry has 39 fields, such as the patient’s ID (same as the national death database), date of birth, gender, town and region of residence, ethnicity, health insurance, length of stay, condition at discharge, and primary and secondary diagnoses according to CDI-10 classification, among others.

#### Inclusion and exclusion criteria

The cohort for our study corresponds to all women who were diagnosed with breast cancer from 2007 to 2018. First, we chose 2007 as the starting year to guarantee that the first discharge in the database is, in fact, the time of diagnosis. Thus, all women with breast cancer discharges from 2007 onward, but none from 2001 to 2007, were included as newly diagnosed patients. Second, we did not include data from 2019 onward because it is not reliable due to the political and social uprising in Chile (October–December 2019) [14] and the COVID-19 pandemic, which largely influenced hospital discharges and deaths. Accordingly, we considered the same period of time for the national death registry.

A discharge and death registry is considered to correspond to breast cancer if its CDI-10 diagnostic code belongs to the categories C50 and D05 (*malignant neoplasm of breast* and *carcinoma in situ of breast*, respectively).

For calculations of mortality rates, we selected all females whose cause of death was directly associated with breast cancer from the 2,549,800 registries in the mortality database, resulting in 16,615 BC deaths after removing 18 death registries with missing IDs, during the study period.

For calculations of incidence rates, we selected all patients whose primary diagnosis was breast cancer from the 32,443,591 hospital discharges, resulting in 154,379 discharges. There were 5,230 deaths that did not have a C50 and D05 discharge in the resulting database including 154,379 discharges. Thus, a set of diagnoses related to breast cancer was considered for such patients (see Appendix A for a detailed explanation), leading to a total of 165,912 breast cancer-related discharges. Then, the cohorts from 2007 until 2018 were selected, resulting in 103,750 records corresponding to 58,914 patients. Out of the 103,750 discharge registries, there were 3,371 with a missing ID. Hence, we do not know if these discharges are associated with new patients or with those already in the cohort of 58,914 patients. Thus, discharge records with missing IDs were removed from the database. However, a sensitivity analysis regarding these registries is performed in the Results section. From the remaining 58,914 patients identified by their IDs, there were 987 patients who had inconsistencies in their gender, i.e.,these patients had hospital discharges where they were identified both as male and female patients. These gender inconsistencies were corrected by considering the gender information from the mortality database and considering the mode of all records associated with such patients. Thus, the gender of 815 people was corrected in the discharges database, while 109 cases for which the mode was inconclusive were deleted. Finally, all female patients were selected, resulting in a total of 99,554 discharge records, corresponding to 58,254 patients, with an average of 1.7 discharges per patient. Figures 1 and 2 summarize the inclusion and exclusion criteria for the death and discharge databases, respectively.

**Figure 1:**
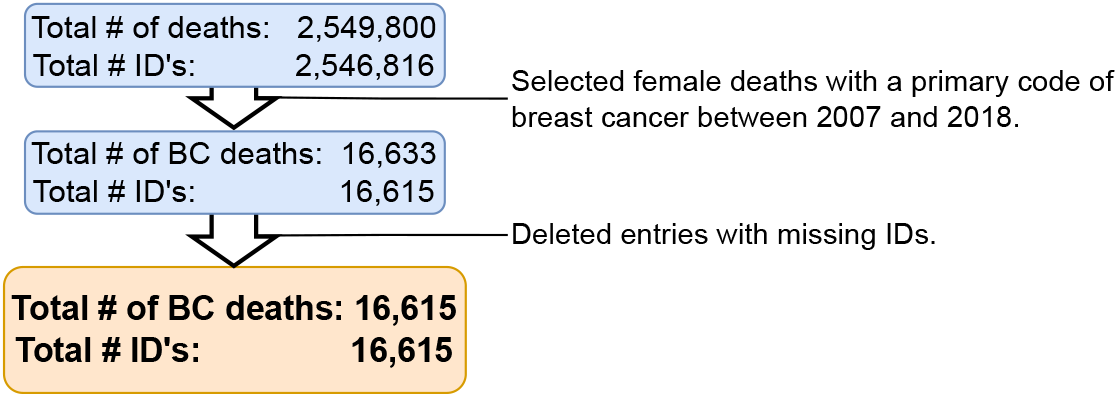
Inclusion and exclusion criteria for the breast cancer death database (2007–2018).

**Figure 2:**
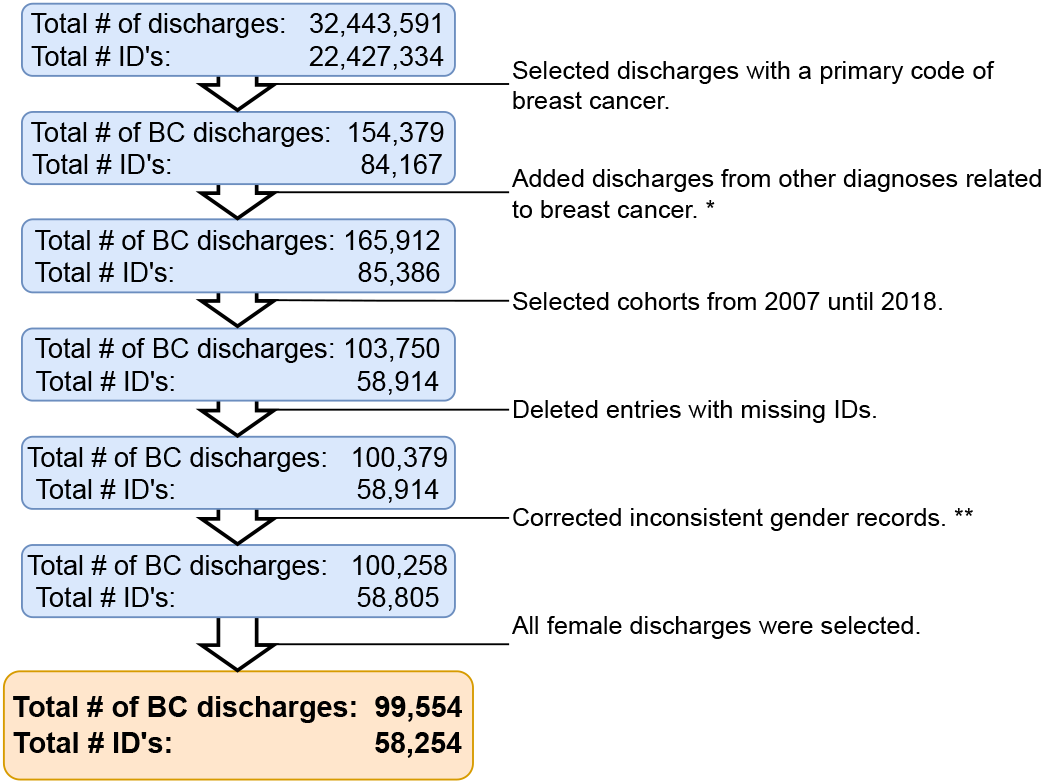
Inclusion and exclusion criteria for the breast cancer hospital discharge database (2007–2018). * Several diagnoses were included considering health problems that could arise due to breast cancer progression or its treatment for patients who died due to breast cancer. See appendix A for further explanation. ** There were patients with records who identified them both as male and female. It was considered that this was an error from the database; therefore, it was corrected.

### 2.2 Methods

We calculated both crude and age adjusted incidence and mortality rates for breast cancer at the national and regional levels. Population estimates and projections provided by the Chilean National Institute of Statistics (INE) [15, 16] were used. For the age-standardized incidence and mortality rates, we used the International Agency for Research on Cancer standard population [17]. We also calculated crude and age-standardized incidence and mortality rates by year, health insurer (public and private systems), and geographical region.

The female population affiliated with the public health system was obtained using the data provided by the FONASA in its yearly statistical bulletin, which is available online from 2009 onward [18]. Prior to this, FONASA bulletins had incomplete information, without segmentation by age intervals or gender. Therefore, we used linear regression to estimate the 2007 and 2008 populations from the available data. Information from the private health system beneficiary population was obtained using the data provided by the Health Superintendence in its yearly statistics publication for ISAPRE’s beneficiaries [19].

Table 1 shows number of deaths, average age of death, and standard deviation by year. The last two columns show the percentage of deaths that do not have a corresponding breast cancer discharge registry, this is, these patients died without hospitalization due to breast cancer, and their average age at death with standard deviation by year. Table 2 shows the number of newly diagnosed breast cancer cases, with the average age of diagnosis and standard deviation.

**Table 1:** Total breast cancer deaths and percentage of breast cancer deaths without breast cancer discharge by year for the period 2007 – 2018. * Women died due to breast cancer, but there were no hospital discharges associated with a breast cancer diagnosis. For the calculation of incidence, it is assumed that these women had a random uniform survival period between 1 and 12 months.

| Year | Total Deaths |  | Deaths without discharge* |  |
| --- | --- | --- | --- | --- |
|  | Cases | Mean Age (std) | Cases(%) | Mean Age (std) |
| 2007 | 1,158 | 65.4 (15.6) | 28.2 | 73.6 (14.8) |
| 2008 | 1,228 | 65.3 (15.6) | 29.2 | 72.8 (15.1) |
| 2009 | 1,337 | 65.8 (15.6) | 24.8 | 75.5 (15.0) |
| 2010 | 1,298 | 64.9 (15.6) | 24.3 | 73.8 (15.3) |
| 2011 | 1,349 | 66.2 (15.5) | 22.4 | 75.2 (15.0) |
| 2012 | 1,371 | 66.4 (15.6) | 23.2 | 74.9 (15.2) |
| 2013 | 1,391 | 65.8 (15.6) | 24.5 | 75.9 (14.7) |
| 2014 | 1,424 | 66.3 (16.0) | 22.9 | 75.9 (15.2) |
| 2015 | 1,512 | 66.5 (15.6) | 23.1 | 76.5 (14.4) |
| 2016 | 1,492 | 66.4 (15.3) | 21.2 | 76.4 (14.7) |
| 2017 | 1,508 | 66.8 (15.3) | 24.3 | 76.7 (14.9) |
| 2018 | 1,547 | 67.1 (16.0) | 23.3 | 77.0 (14.5) |
| Total | 16,615 | 66.1 (15.6) | 24.1 | 75.4 (14.9) |

**Table 2:** New breast cancer cases by year for 2007 – 2018.

| Year | New cases | Mean Age (std) |
| --- | --- | --- |
| 2007 | 3,785 | 57.4 (14.1) |
| 2008 | 3,938 | 57.5 (14.1) |
| 2009 | 4,576 | 57.3 (13.9) |
| 2010 | 4,415 | 57.9 (14.1) |
| 2011 | 4,685 | 57.7 (14.0) |
| 2012 | 4,995 | 57.3 (13.8) |
| 2013 | 5,172 | 57.7 (13.7) |
| 2014 | 4,932 | 58.1 (13.7) |
| 2015 | 5,427 | 57.8 (13.6) |
| 2016 | 5,569 | 57.5 (13.8) |
| 2017 | 5,325 | 58.1 (14.0) |
| 2018 | 5,435 | 58.1 (13.9) |
| Total | 58,254 | 57.7 (13.9) |

## 3 Results

The number of new breast cancer diagnoses increased by 43.6%, from 3,785 in 2007 to 5,435 in 2018. Total breast cancer deaths increased by 33.6% from 1,158 to 1,547 during the same period of time. We noticed a slight increasing trend in the mean age at death, with a 2.6% growth from 65.4 to 67.1, while there was no significant change in the mean age at diagnosis. We noted that the female population in Chile grew 13.3% from 8,390,194 to 9,506,921 during the period 2007–2018. According to these records, each woman had an average of 1.67 hospital discharges per patient. In what follows, the results of incidence and mortality rates are shown.

### 3.1 Incidence rates

As explained in Subsection 2.2, incidence rates are calculated based on a set of 62,093 patients. In total, 58,254 patients were identified through the discharge registries plus the 3,839 breast cancer death registries without a discharge registry. Figure 3 shows both crude and age–adjusted incidence rates (cases/100,000 women). We observe that the crude incidence rates are higher than the age–adjusted rates and have a growing trend over time, with an increase of 19.7%, from 49.2 in 2007 to 58.9 in 2018. The age–adjusted rates are stable over time, with an average rate of 44.0 and a standard deviation of 2.2 during the study period.

**Figure 3:**
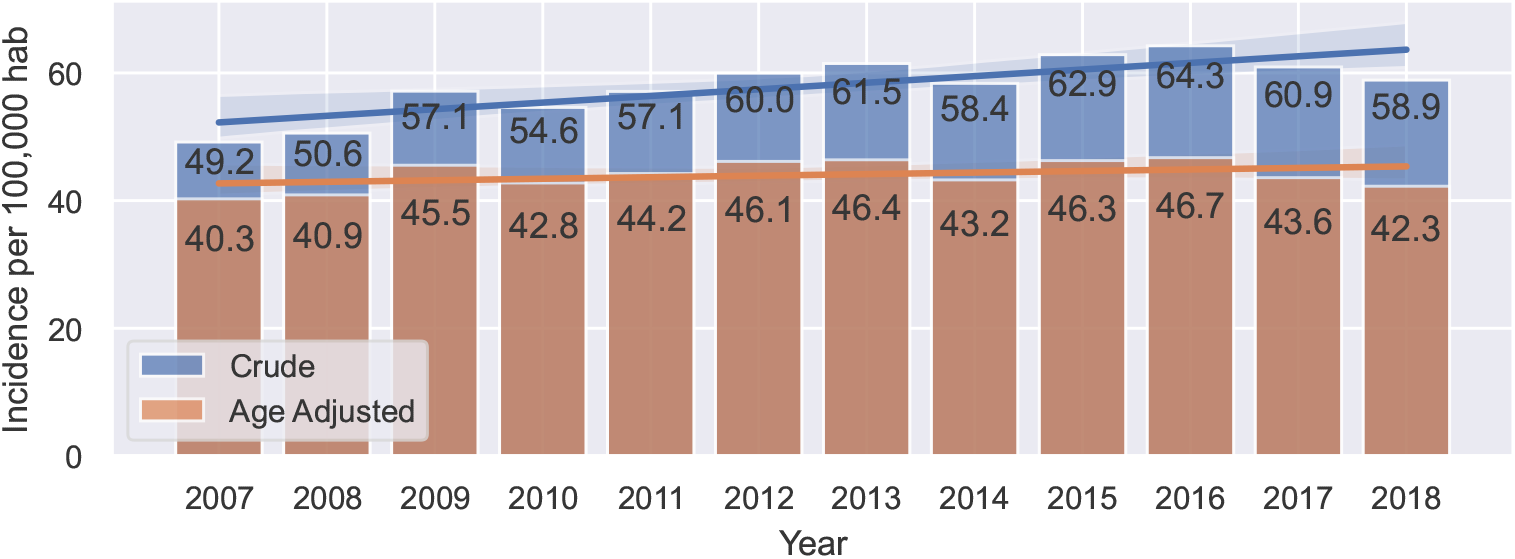
Crude and age–adjusted incidence rates by year (cases/100,000 women).

It is important to note that there were 3,371 discharge registries that were deleted for having no ID and, therefore, it cannot be determined if these registries correspond to patients who have been already identified in the cohort or if they correspond to new diagnoses. To evaluate the impact of these cases on the incidence estimates, we considered two possible scenarios. First, we considered the worst–case scenario, where all 3,371 registries are new breast cancer diagnoses. Table 3 shows the crude incidence rates calculated, accounting for each of these registries as a new diagnosis. Table 4 shows a more likely scenario, where some of these discharges do not belong to a new patient. Women from the resulting database had an average of 1.67 hospital discharges; then, as a more likely scenario, we considered that there is a newly diagnosed patient for every 1.67 discharges with missing IDs.

**Table 3:** Registries with missing IDs and crude incidence rates (cases/100,000 women) calculated considering these as new cases (worst case scenario). The last row shows the variation between these crude rates and those shown in Figure 3.

| Year | 2007 | 2008 | 2009 | 2010 | 2011 | 2012 | 2013 | 2014 | 2015 | 2016 | 2017 | 2018 |
| --- | --- | --- | --- | --- | --- | --- | --- | --- | --- | --- | --- | --- |
| Registries with missing IDs | 1286.0 | 528.0 | 276.0 | 241.0 | 225.0 | 245.0 | 206.0 | 164.0 | 28.0 | 28.0 | 47.0 | 97.0 |
| Crude Incidence | 64.5 | 56.8 | 60.4 | 57.4 | 59.7 | 62.7 | 63.8 | 60.2 | 63.2 | 64.6 | 61.4 | 59.9 |
| Variation (%) | 23.8 | 11.0 | 5.3 | 4.8 | 4.3 | 4.4 | 3.6 | 3.0 | 0.5 | 0.5 | 0.8 | 1.7 |

**Table 4:** Registries with missing IDs and crude incidence rates (cases/100,000 women) calculated considering only a portion of these as new cases (likely scenario). This equals one new diagnosis every 1.67 discharges. The last row shows the variation among these crude rates and those shown in Figure 3.

| Year | 2007 | 2008 | 2009 | 2010 | 2011 | 2012 | 2013 | 2014 | 2015 | 2016 | 2017 | 2018 |
| --- | --- | --- | --- | --- | --- | --- | --- | --- | --- | --- | --- | --- |
| Registries with missing IDs | 772.0 | 317.0 | 166.0 | 145.0 | 135.0 | 147.0 | 124.0 | 98.0 | 17.0 | 17.0 | 28.0 | 58.0 |
| Crude Incidence | 58.4 | 54.3 | 59.1 | 56.3 | 58.6 | 61.6 | 62.9 | 59.5 | 63.1 | 64.4 | 61.2 | 59.5 |
| Variation (%) | 15.8 | 6.9 | 3.3 | 3.0 | 2.6 | 2.7 | 2.2 | 1.8 | 0.3 | 0.3 | 0.5 | 1.0 |

There was a large number of registries with missing IDs in 2007, accounting for more than a third of all registries with missing IDs. The years prior to 2007 also show a large number of discharges with missing IDs, with a yearly average of 1,820 for the 2001-2006 period. This may be due to registration errors during the first years that were corrected, resulting in a much lower number of missing IDs in the later years of the period.

As expected, the crude incidence rates obtained in both cases are higher than those shown in Figure 3, with the highest differences in 2007 and 2008. In the worst–case scenario, apart from these two years, all variations are lower than 5.3%. The mean crude rate from 2007–2018 was 61.2/100,000 women, with a standard deviation of 2.6. The mean crude incidence rate previously found is 57.9/100,000 women, with a variation between the two crude rates of 5.3%, which represents an upper bound considering the worst–case scenario for the error on the estimates shown in Figure 3. In the more likely scenario shown in Table 4, in addition to 2007 and 2008, all crude rates have a variation lower than 3.3%. The mean crude rate from 2007–2018 was 59.9/100,000 women, with a standard deviation of 2.9. This represents a variation with respect to the mean crude rate found in Figure 3 of 3.3%, which represents an upper bound considering a more likely scenario for the error.

There are considerable differences in age–adjusted incidence rates among regions, as shown in Figure 4, with no clear geographical trend. Higher incidence rates are found in the northern part of the country, the Arica and Parinacota region (XV), and in the central part of the country, corresponding to the Metropolitan region (RM) and Valparaiso region (V), with age–standardized incidence rates of 50.9, 46.8 and 45.5, respectively. Interestingly, two of the regions with the lowest rates were also found in the northern area, the Atacama (III) and Tarapacá (I) regions, with incidences of 28.2 and 28.3, respectively, surpassed only by a southern and a central region, Los Lagos (X) and O’Higgins (VI), with age–adjusted incidence rates of 26.5 and 27.3, respectively. The rest of the regions have an age–adjusted rate between 30.0 and 42.7.

**Figure 4:**
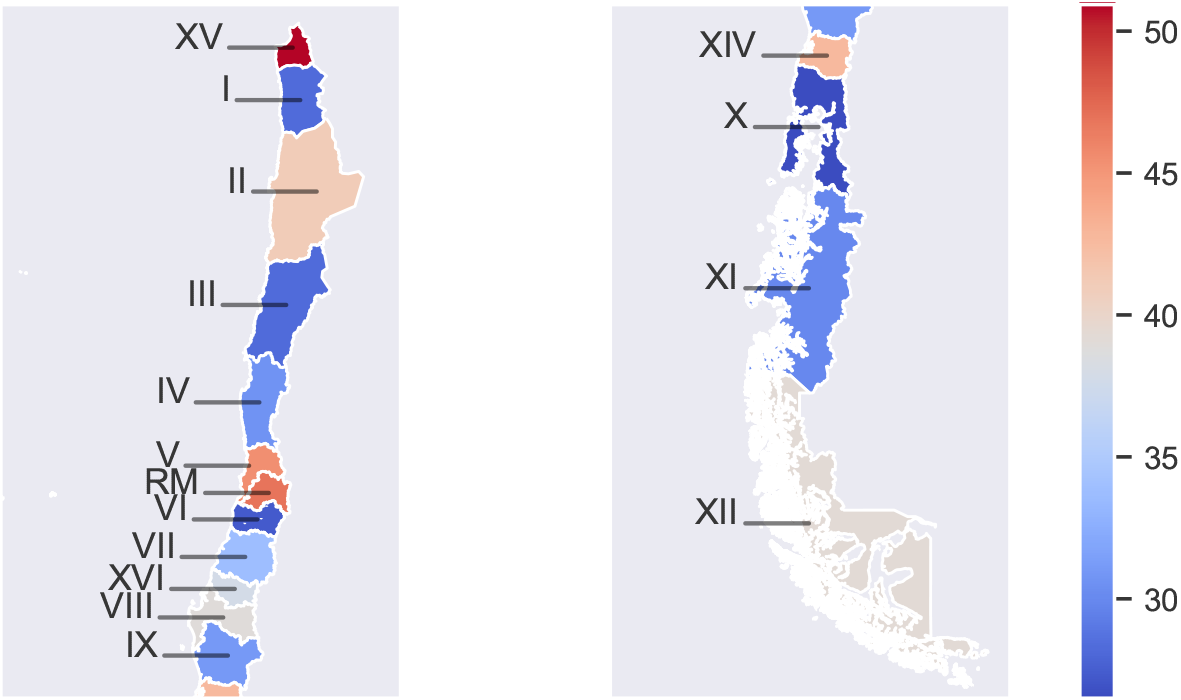
Mean age–adjusted incidence rates (cases/100,000 women) during 2007-2018 by geographical region. Detailed incidence values can be found in Table B.1.

Table 5 shows crude and age adjusted incidence rates by year for women affiliated with the private and public health systems. For this analysis, 56,250 out of the 62,093 registries were considered, i.e., a total of 90.6% of all diagnosed women in the 2007–2018 period had an affiliation with the private and public health systems. Of the remaining 5,843 women, 1,944 women had armed forces health insurance, 885 women had no health insurance, and 3,014 women had missing information.

**Table 5:** Crude and age–adjusted incidence rates (cases/100,000 women) for women with private and public health insurance.

| Year | ISAPRE |  | FONASA |  |
| --- | --- | --- | --- | --- |
|  | Crude | Age adjusted | Crude | Age adjusted |
| 2007 | 36.3 | 37.2 | 46.7 | 36.7 |
| 2008 | 41.5 | 41.9 | 49.1 | 37.9 |
| 2009 | 72.3 | 71.1 | 54.9 | 42.9 |
| 2010 | 66.0 | 65.3 | 50.5 | 37.4 |
| 2011 | 61.1 | 58.4 | 54.2 | 39.2 |
| 2012 | 64.2 | 61.2 | 56.4 | 40.6 |
| 2013 | 63.1 | 59.2 | 57.8 | 40.6 |
| 2014 | 71.3 | 66.5 | 53.3 | 36.5 |
| 2015 | 72.9 | 67.0 | 59.1 | 40.7 |
| 2016 | 84.0 | 75.1 | 57.6 | 39.3 |
| 2017 | 73.0 | 62.8 | 55.2 | 37.5 |
| 2018 | 72.3 | 61.2 | 53.6 | 36.7 |
| Mean (Std) | 64.8 (13.6) | 60.6 (11.0) | 54.0 (3.7) | 38.8 (2.0) |

We observed that women affiliated with a private provider (ISAPRE) had higher incidence rates during the period under study, having an average age–adjusted incidence rate of 60.6/100,000 compared to 38.8/100,000 for women affiliated with the public provider (FONASA). The age–standardized incidence rate for the private health system shows greater fluctuations over time (sd of 11.0) than that for the public sector (sd of 2.0).

### 3.2 Mortality rates

Figure 5 shows both crude and age–adjusted mortality rates (deaths/100,000 women) for the period under study. Crude mortality rates increased 18.1%, from 13.8 per 100,000 women in 2007 to 16.3 per 100,000 women in 2018. On the other hand, age–adjusted mortality rates did not have significant changes during the study period, with an average standardized mortality rate of 10.5 per 100,000 women and a standard deviation of 0.4.

**Figure 5:**
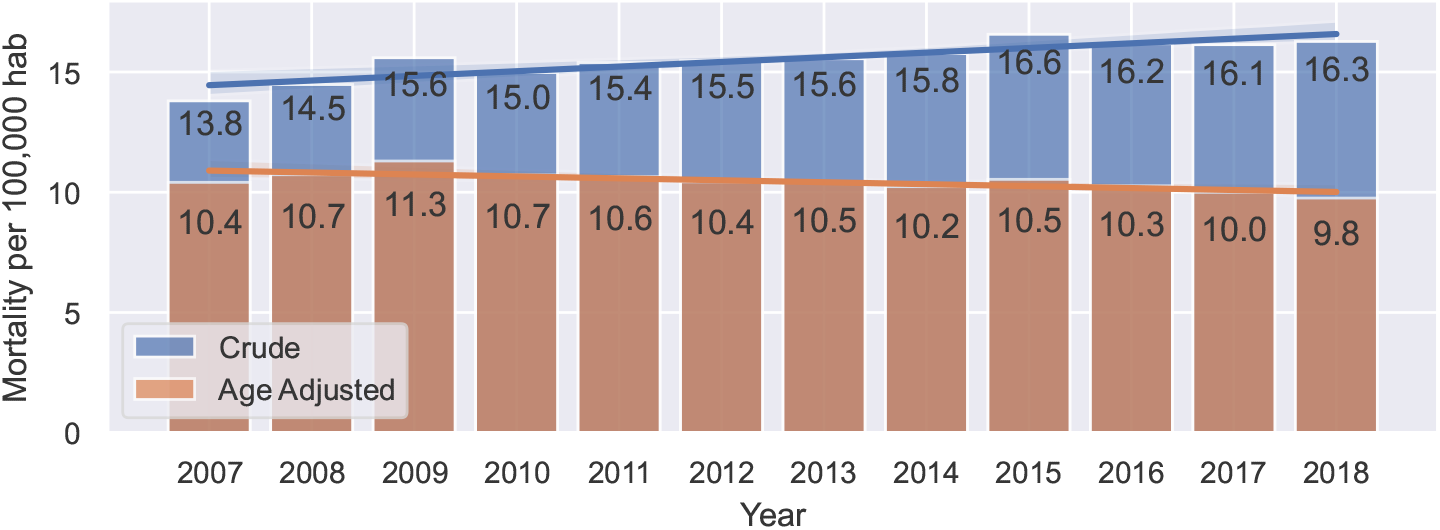
Crude and age–adjusted mortality rates (deaths/100,000 women) by year.

Figure 6 shows that the southern cities of Magallanes and Antártica Chilena (XII) have the highest age–adjusted mortality rate of 13.1, followed by the Valparaiso (V), Ñuble (XVI) and Metropolitan (RM) regions, with average age–adjusted rates of 11.7, 10.9, and 10.8, respectively. On the other hand, the regions with the lowest mortality rates are the southern regions of Los Lagos (X) and Los Ríos (XIV), with age–adjusted mortality rates of 8.1 and 8.9, respectively. The rest of the regions have similar mortality rates, averaging between 9.0 and 10.6 deaths per 100,000 women.

**Figure 6:**
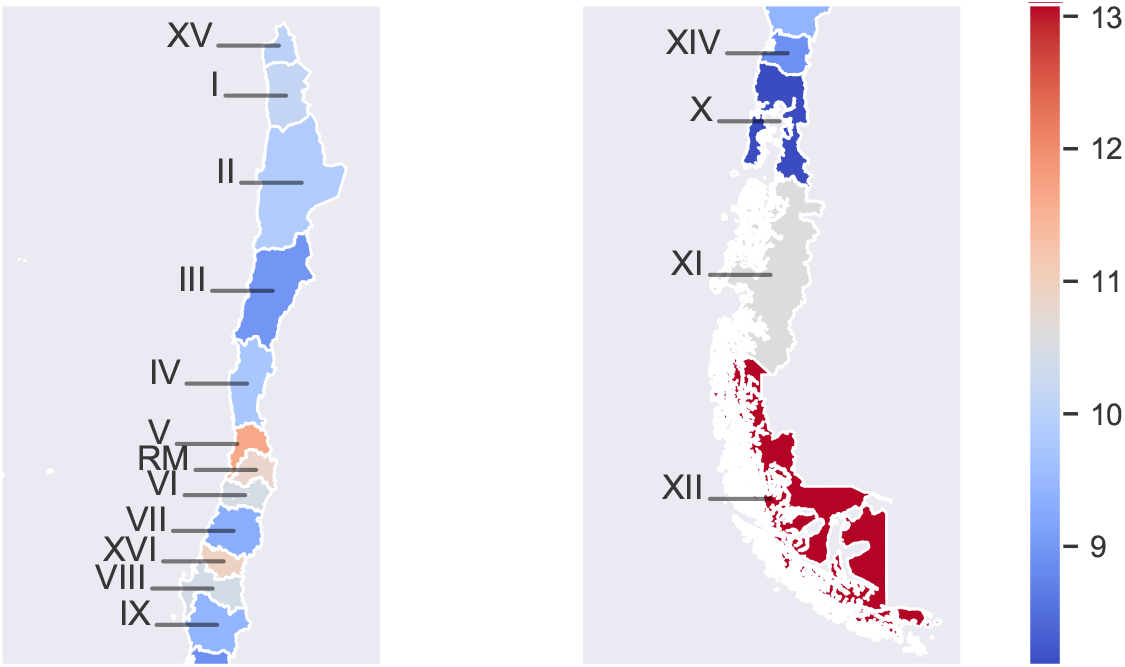
Mean age–adjusted mortality rates (deaths/100,000 women) during 2002-2018 by geographical region. Detailed mortality values can be found in Table B.1.

Table 6 shows mortality rates by year for women affiliated with the private and public health systems. As the national death registry does not include the health insurance of the deceased patients, we obtained them by searching the discharge database with the corresponding patient ID. Information was found for 89.3% of all women from the death database, corresponding to 14,837 women, though the rest did not have any hospital discharge data. From this set of women, 14,057 women belong to either the FONASA or ISAPRE. The last column of Table 6 shows the percentage of women from the death database from which it was not possible to recover their health care provider.

**Table 6:** Crude and age–adjusted mortality rates (deaths/100,000 women) for women with private and public health insurance. The last columns show the percentage of registries with no health insurance data.

| Year | ISAPRE |  | FONASA |  | Missing (%) |
| --- | --- | --- | --- | --- | --- |
|  | Crude | Age adjusted | Crude | Age adjusted |  |
| 2007 | 8.4 | 9.8 | 12.4 | 9.0 | 18.0 |
| 2008 | 8.0 | 8.6 | 13.0 | 9.2 | 16.9 |
| 2009 | 9.2 | 10.3 | 15.4 | 11.0 | 13.1 |
| 2010 | 9.4 | 10.2 | 13.9 | 9.5 | 13.3 |
| 2011 | 9.8 | 10.5 | 14.7 | 9.5 | 10.8 |
| 2012 | 8.7 | 8.8 | 15.0 | 9.3 | 9.5 |
| 2013 | 10.1 | 10.6 | 15.0 | 9.5 | 8.4 |
| 2014 | 9.7 | 10.1 | 15.5 | 9.4 | 8.7 |
| 2015 | 11.7 | 11.9 | 16.1 | 9.6 | 9.3 |
| 2016 | 10.3 | 9.8 | 16.1 | 9.6 | 7.7 |
| 2017 | 11.3 | 10.5 | 15.5 | 9.2 | 8.0 |
| 2018 | 11.1 | 9.8 | 15.5 | 8.9 | 8.3 |
| Mean (Std) | 9.8 (1.2) | 10.1 (0.9) | 14.8 (1.2) | 9.5 (0.5) | 10.7 |

We observe, in terms of crude rates, that FONASA patients have a higher mortality rate than ISAPRE patients, with rates of 14.8 and 9.8 per 100,000 women, respectively. When adjusted by age groups, FONASA rates decreased, while ISAPRE rates increased, resulting in FONASA patients having a slightly lower age–adjusted mortality rate of 9.5 compared to 10.1 for ISAPRE patients. Both systems had a rather constant trend in terms of age–standardized rates throughout the study period.

## 4 Discussion

Breast cancer is one of the two cancers, together with gallbladder cancer [20], that most frequently affects Chilean women. It is also the leading cause of death from cancer for Chilean women [21]. In 1995, the Cancer Unit of the Ministry of Health formed the National Breast Cancer Commission to focus on the design of a National Breast Cancer Program to better organize the care for breast diseases [22]. The commission defined referral, diagnostic, and treatment guidelines and carried out several activities across the country to improve breast cancer patients’ care. As of 2004, a major reform in the Chilean health system was introduced: the Universal Access Plan for Explicit Guarantees (AUGE), later renamed GES, which aimed to provide timely and quality health care without discrimination and with financial protection to all Chilean residents. Thus, the Ministry of Health, through this plan, guarantees the coverage of a number of diseases (It started in 2005 with 25 pathologies and grew to 85 in 2022). Breast cancer was part of the initial pathologies included in 2005; in addition to the quality, access, financial, and opportunity guarantees, it also defined the complete protocol of medical treatments, including procedures and drugs to diagnose and treat breast cancer in all its stages. This protocol was updated in 2010 and 2015 according to the new treatment options that had been developed over time. In particular, in 2015, the high–cost drug trastuzumab was included for all Chilean women with HER2–positive breast cancer. The GES plan activates the above guarantees at the moment of a suspicion of breast cancer for people aged 15 years and over with suspected, diagnosed or recurrent breast cancer [23]).

Regarding early breast cancer detection, the Ministry of Health currently *recommends* an annual mammogram for women 50 to 69 years old (www.diprece.minsal.cl). However, despite this recommendation, it only guarantees triannual screening mammograms free of charge for all women 50 to 59 years old, independent of their health insurer [23]. The National Socioeconomic Characterization Survey, CASEN 2017 [24], shows that in 2017, only 53.4% and 33.1% of women aged 50 or more in the private and public health systems had a screening mammogram during the last year, respectively. These figures increase to 76.3% and 57.1% for a screening mammogram in the last three years.

These results might have changed with the implementation, as of July 2019, of a MINSAL strategy of increasing the coverage of mammograms in public primary health care, with important investments in new mammographers and the hiring of human resources in Family Health Centers (CESFAM) and community hospitals. One specific goal of this plan was to improve the coverage of breast cancer screening mammograms. For this purpose, the exams are taken by medical technologists in the aforementioned health care facilities and are sent electronically to the *mammography cell* (digital hospital), where the report is made by radiologists specialized in breast imaging, totaling 22% of the mammography reports in the country from January to September 2021 [25].

However, we note that in Chile there is no screening program as defined by the World Health Organization [26]. In particular, there is no information system in place linked to population registries that would allow the screenings to be offered to the eligible population using a call and recall system and have a registry of all screenings performed in the country.

With regard to epidemiological indicators, Chile does not have a national cancer registry. Hence, key estimates, such as incidence and case fatality rates, have been extrapolated from population records created in 1998 and updated until 2012 in the regions of Antofagasta, Biobío, Valdivia and Concepción [27]. These regional data are insufficient to have a comprehensive view of the evolution of cancer at the national level due to differences in health care coverage, ethnicity, and regional particularities, among others. Furthermore, there is no reliable information from 2012 onward. Thus, we observe that most of the published research focuses on mortality indicators, of which there is a reliable national death database. However, regarding incidence rates, there are few studies with assumptions that might lead to inaccurate estimates. For example, the MINSAL study in 2019 [3] reported a constant drop in incidence rates, which is inconsistent with the data of the current state of breast cancer in Chile. This drop is obtained by the actual decrease in mortality rates and the assumption of a constant fatality rate over the study period, without considering the improvements in breast cancer treatments over time. These estimates feed the Chilean data in GLOBOCAN [5].

The incidence age-standardized rates found for the 2007–2018 period are lower than crude rates for the same period. This is largely explained by the demographic composition of the Chilean population. Breast cancer worldwide is significantly more common in older women and is less likely in women younger than 30 years old. This distribution is also observed in Chile. As Chile has a higher proportion of older women than the standardized population [15, 17], it is expected to have higher crude incidence rates. A similar result is found for mortality rates in the 2007–2018 period. The Global Cancer Observatory (GLOBOCAN) reported the Chilean incidence and mortality age–adjusted rates as 37.4 and 10.2 per 100,000 women in 2020, respectively [2]. Our study estimates incidence and mortality age adjusted rates as 44.0 and 10.5 per 100,000 women, respectively, with a constant trend over the 2007–2008 period. We observe that our estimates of the mortality rate are consistent with those reported by GLOBOCAN, while our estimation of the incidence rate is higher than that of GLOBOCAN. The trends in age–standardized incidence reported by GLOBOCAN show that almost all countries have either had a constant trend (in countries such as Canada, Italy and Poland) or a growing trend (in countries such as Germany, Thailand or Korea) in the last 20 years. We remark that the constant trend found in our study for age–standardized incidence rates at the national level is consistent with this global trend observed by GLOBOCAN and differs from the decreasing trend found by the Ministry of Health [3]. It is important to note that the GLOBOCAN platform reports data on incidence and mortality based on information provided by countries and has no responsibility over their quality [28].

We also observe from Table 5 a much higher age–adjusted incidence rate in privately insured patients (ISAPRE) compared to those with public health care insurance. As discussed in the Introduction, these two groups of patients have important differences, such as income, lifestyle, diet composition, comorbidities and use of hormonal therapies, among others, which might explain these incidence differences [29]. Another plausible explanatory hypothesis for this difference might rely on the higher use of spontaneous screening due to better access and awareness of its importance among privately insured women [24] and on the other hand, women who are public system beneficiaries are not well–informed about their right of having every 3 years a free mammogram between 50 to 59 years-old, which should increase the detection at early stages and at early ages, or even in older women who otherwise would not be tested [30].

The estimated incidence and mortality rates shown by region in Figures 4 and 6 show the great differences in breast cancer epidemiology along the Chilean territory. These differences indicate that the reality of each region must be taken into account when designing public health measures. Moreover, public measures and other related policies may be misdirected if they are designed and evaluated only through a few local registries, such as those from Antofagasta, Biobío and Los Rios regions. This highlights the need for a national–level cancer registry from which adequate data can be obtained.

### Study strength and limitations

To the best of our knowledge, this is the first study on breast cancer incidence rates at the national level using publicly available data from several national registries on hospital discharges. Incidence has been reported by GLOBOCAN, estimating a 2020 incidence rate of 37.4. GLOBOCAN’s estimate is lower than the estimation of this study, and even more so considering the constant trend shown in Figure 3, with an average incidence of 42.3. Furthermore, this study highlights the important differences in key health care indicators for women across regions, age, and health care insurance plans. The study of case fatality and survival rates for different segments of Chilean women is part of our ongoing research as well as the causes of these differences.

This study also has some limitations. First is the limitation of the quality of the national database used for hospital discharges and deaths. As discussed previously, there are missing and conflicting data and ambiguity in the cause of death in some death certificates. Furthermore, we used a hospital discharge database, and therefore, there might be some patients diagnosed with breast cancer who are never hospitalized when receiving treatment (for example, those who do not have surgery). Thus, incidence rates might be underestimated.

## 5 Conclusions

The methodology used in this study presents an alternative for obtaining incidence and mortality rates from public databases, which generates results consistent with other measures, specially in the absence of a national cancer registry, such is the case in Chile. We remark however, that, with the purpose of developing, implementing, and evaluating cancer public policies, it is of the utmost importance and priority to have a national cancer registry in Chile.

## Data Availability

Two public anonymized databases were used in this study, these are the National Death Registry and the Hospitalary Discharge Registry. Both datasets were provided by the Chilean Ministry of Health through the Department of Health Statistics and Information following the Data Transparency Law. This data is available in their open data website https://deis.minsal.cl/#datosabiertos.

## Data Availability

Scientists can obtain access to individual-level data from the UK Biobank by applying to UK Biobank (https://www.ukbiobank.ac.uk/enable-your-research).

## Acknowledgement

The authors gratefully acknowledge financial support from ANID PIA/APOYO AFB180003 and CMM-Conicyt PIA AFB170001.

## Appendix A Other breast cancer diagnoses

After selecting discharges with a primary diagnosis code of breast cancer, there were still a considerable number of breast cancer death registries that did not have any associated discharge registry (5,230 deaths). However, there are discharge registries for causes other than breast cancer that are not typically included in a breast cancer incidence analysis. These are health problems that can arise due to the progression of the disease or its treatment (surgery, chemotherapy, radiotherapy) and therefore will be included under certain specific conditions. Thus, these diagnostics were taken into consideration only when they belonged to a patient who died because of breast cancer. We group these diagnostics into three groups:

1. Discharge registries directly attributable to breast cancer and its treatment. This item includes examination, treatment by chemotherapy and radiotherapy, and different breast–related diseases and issues, which may be due to breast cancer misdiagnosis. These diagnostics are D486, D24X, Z123, Z803, Z853, Z031, Z080, Z081, Z082, Z087, Z088, Z089, Z129, Z400, Z510, Z511, Z512, Z515, Z809, Z859, and Z860.
2. Diagnostics of other cancers and malignancies associated with breast cancer. This item includes secondary tumors and tumors of unspecified places. These diagnostics are C798, C782, C795, C793, C787, C786, C412, C800, D382, C792, C709, C383, C799, D059, C770, D420, C796, C414, C500, C413, C771, C728, C967, C779, D383, C399, C773, C781, C783, and C700.
3. Other diagnostics might be attributable to symptoms of breast cancer and its treatment. They are considered only if such discharge is close enough to the death registry. Each diagnosis has a different period of time to be associated with death and is shown in Table A.1. The absence of a period means that the diagnosis is always included independent of its gap from the death registry.

**Table A.1:** CDI-10 codes and relation periods of the health problems to be considered as the breast cancer debut.

| Diagnostic Code | Relation period (years) | Diagnostic Code | Relation period (years) |
| --- | --- | --- | --- |
| D649 | 2 | N850 | - |
| G039 | 1 | R060 | 1 |
| G540 | 2 | R17X | 1 |
| G939 | 1 | R18X | 1 |
| G952 | 2 | R51X | 1 |
| I495 | - | R53X | 1 |
| I891 | 1 | S220 | 4 |
| I972 | - | S320 | 2 |
| J80X | 2 | S323 | 4 |
| J90X | 2 | S325 | 4 |
| J948 | 2 | S327 | 4 |
| J960 | 2 | S328 | 4 |
| J969 | 1 | S423 | 4 |
| J984 | 2 | S720 | 4 |
| J989 | 2 | S721 | 4 |
| M532 | 4 | S722 | 4 |
| M544 | 4 | S723 | 4 |
| M546 | 4 | S724 | 4 |
| M549 | 4 | S728 | 4 |
| M808 | 4 | S729 | 4 |
| M844 | 4 | T08X | 4 |
| N63X | - | T12X | 4 |
| N645 | - | T142 | 4 |
| N648 | - | T932 | 4 |
| N649 | - |  |  |

Through the addition of these diagnoses, 11,533 discharge registries were added to the discharge database, which corresponds to 1,219 patients.

## Appendix B Incidence and Mortality Results

### B.1 Breast cancer incidence and mortality by geographical region

In table B.1 displays the mean incidence over the period 2007–2018 and mortality over the period 2007–2018 for each of the 16 Chilean regions. Both are presented as age–adjusted and crude rates. They are sorted from north to south.

**Table B.1:**
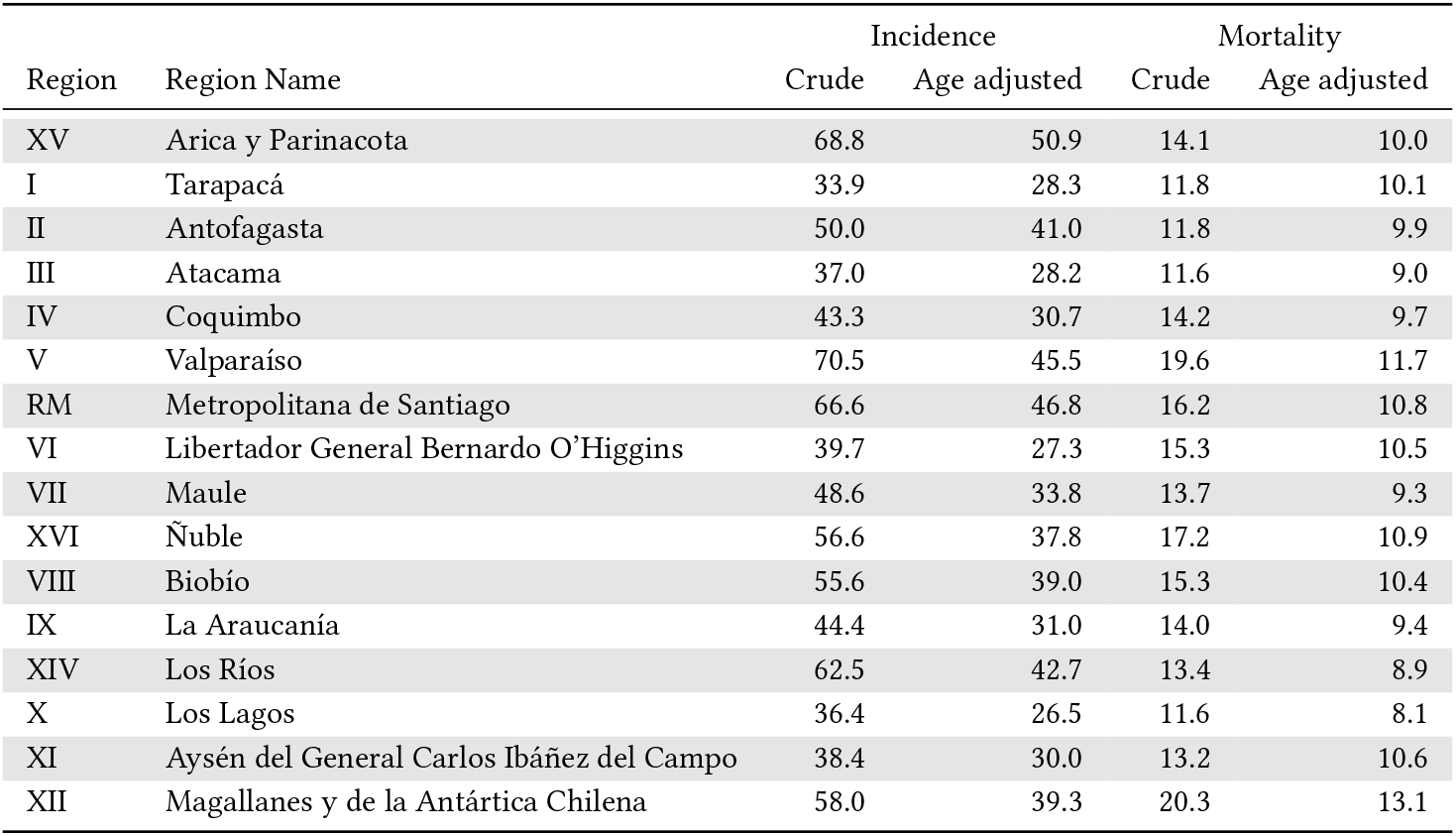
Average incidence and mortality (cases/100,000 women) over the period 2007–2018 by region.

| Region | Region Name | Incidence |  | Mortality |  |
| --- | --- | --- | --- | --- | --- |
|  |  | Crude | Age adjusted | Crude | Age adjusted |
| XV | Arica y Parinacota | 68.8 | 50.9 | 14.1 | 10.0 |
| I | Tarapacá | 33.9 | 28.3 | 11.8 | 10.1 |
| II | Antofagasta | 50.0 | 41.0 | 11.8 | 9.9 |
| III | Atacama | 37.0 | 28.2 | 11.6 | 9.0 |
| IV | Coquimbo | 43.3 | 30.7 | 14.2 | 9.7 |
| V | Valparaíso | 70.5 | 45.5 | 19.6 | 11.7 |
| RM | Metropolitana de Santiago | 66.6 | 46.8 | 16.2 | 10.8 |
| VI | Libertador General Bernardo O'Higgins | 39.7 | 27.3 | 15.3 | 10.5 |
| VII | Maule | 48.6 | 33.8 | 13.7 | 9.3 |
| XVI | Ñuble | 56.6 | 37.8 | 17.2 | 10.9 |
| VIII | Biobío | 55.6 | 39.0 | 15.3 | 10.4 |
| IX | La Araucanía | 44.4 | 31.0 | 14.0 | 9.4 |
| XIV | Los Ríos | 62.5 | 42.7 | 13.4 | 8.9 |
| X | Los Lagos | 36.4 | 26.5 | 11.6 | 8.1 |
| XI | Aysén del General Carlos Ibáñez del Campo | 38.4 | 30.0 | 13.2 | 10.6 |
| XII | Magallanes y de la Antártica Chilena | 58.0 | 39.3 | 20.3 | 13.1 |

### B.2 Breast cancer incidence by age group

Table B.2 presents age–specific incidence rates for each age interval for each year from 2007 to 2018.

**Table B.2:** Crude incidence rate per year and age group (cases/100,000 women).

| Age | 2007 | 2008 | 2009 | 2010 | 2011 | 2012 | 2013 | 2014 | 2015 | 2016 | 2017 | 2018 | Mean (Std) |
| --- | --- | --- | --- | --- | --- | --- | --- | --- | --- | --- | --- | --- | --- |
| 0-19 | 0.6 | 0.6 | 0.7 | 0.8 | 0.6 | 0.7 | 0.8 | 0.6 | 0.8 | 0.5 | 0.7 | 0.5 | 0.7 (0.1) |
| 20-24 | 2.0 | 3.2 | 3.8 | 2.3 | 3.3 | 3.0 | 3.8 | 2.0 | 2.2 | 3.5 | 4.5 | 3.1 | 3.1 (0.8) |
| 25-29 | 7.1 | 8.0 | 5.1 | 7.7 | 6.1 | 5.6 | 6.5 | 5.8 | 6.4 | 8.5 | 6.8 | 8.2 | 6.8 (1.0) |
| 30-34 | 11.6 | 12.1 | 14.0 | 14.0 | 13.0 | 18.9 | 16.0 | 15.2 | 17.0 | 20.5 | 17.8 | 17.9 | 15.7 (2.7) |
| 35-39 | 29.1 | 28.6 | 36.5 | 30.9 | 35.2 | 36.7 | 36.9 | 34.6 | 38.8 | 36.6 | 36.0 | 35.7 | 34.6 (3.1) |
| 40-44 | 60.5 | 61.2 | 66.9 | 59.7 | 67.1 | 74.4 | 72.9 | 68.1 | 70.5 | 80.3 | 72.1 | 65.2 | 68.2 (5.9) |
| 45-49 | 96.7 | 92.8 | 111.3 | 102.4 | 111.5 | 117.7 | 108.3 | 103.0 | 113.7 | 121.7 | 98.7 | 105.9 | 107.0 (8.3) |
| 50-54 | 104.2 | 108.1 | 125.1 | 113.5 | 113.7 | 125.0 | 122.2 | 110.0 | 133.0 | 130.7 | 108.5 | 118.0 | 117.7 (9.1) |
| 55-59 | 121.6 | 113.3 | 140.7 | 120.7 | 122.1 | 122.4 | 140.9 | 131.4 | 142.1 | 130.9 | 131.0 | 124.9 | 128.5 (8.9) |
| 60-64 | 150.6 | 156.5 | 164.7 | 160.0 | 161.1 | 164.7 | 158.3 | 146.3 | 142.9 | 143.7 | 146.6 | 138.9 | 152.9 (8.7) |
| 65-69 | 160.1 | 179.5 | 172.1 | 172.4 | 182.6 | 186.0 | 197.3 | 175.4 | 181.4 | 170.5 | 174.6 | 150.6 | 175.2 (11.5) |
| 70-74 | 147.3 | 145.8 | 175.1 | 181.8 | 177.8 | 179.8 | 187.7 | 195.8 | 197.0 | 188.1 | 187.8 | 186.8 | 179.2 (15.9) |
| 75-79 | 154.1 | 164.4 | 176.3 | 160.7 | 172.3 | 178.3 | 172.8 | 165.8 | 190.1 | 198.6 | 182.6 | 171.4 | 174.0 (11.9) |
| 80-84 | 178.6 | 163.7 | 164.9 | 189.2 | 168.8 | 163.1 | 179.7 | 172.2 | 165.8 | 168.2 | 148.5 | 143.6 | 167.2 (12.0) |
| 85+ | 235.9 | 236.0 | 228.9 | 206.2 | 209.6 | 172.6 | 178.4 | 171.4 | 147.7 | 165.6 | 171.2 | 107.0 | 185.9 (37.2) |

### B.3 Breast cancer mortality by age group

Table B.3 presents age–specific mortality rates for each age interval for each year from 2007 to 2018.

**Table B.3:** Crude mortality rate by year and age group (cases/100,000 women).

| Age | 2007 | 2008 | 2009 | 2010 | 2011 | 2012 | 2013 | 2014 | 2015 | 2016 | 2017 | 2018 | Mean (Std) |
| --- | --- | --- | --- | --- | --- | --- | --- | --- | --- | --- | --- | --- | --- |
| 0-19 | 0.0 | 0.0 | 0.0 | 0.0 | 0.0 | 0.0 | 0.0 | 0.0 | 0.0 | 0.0 | 0.0 | 0.0 | 0.0 (0.0) |
| 20-24 | 0.0 | 0.0 | 0.1 | 0.1 | 0.0 | 0.1 | 0.0 | 0.0 | 0.1 | 0.0 | 0.1 | 0.1 | 0.1 (0.1) |
| 25-29 | 0.5 | 1.2 | 0.9 | 0.9 | 1.1 | 0.3 | 1.2 | 0.9 | 0.4 | 0.5 | 0.3 | 1.0 | 0.8 (0.3) |
| 30-34 | 1.1 | 3.0 | 1.4 | 2.0 | 2.0 | 1.9 | 2.0 | 2.1 | 2.6 | 2.1 | 2.9 | 3.4 | 2.2 (0.6) |
| 35-39 | 4.6 | 5.3 | 5.5 | 6.0 | 4.0 | 4.9 | 5.0 | 7.0 | 6.2 | 4.8 | 4.1 | 5.2 | 5.2 (0.8) |
| 40-44 | 11.1 | 9.3 | 9.7 | 11.4 | 9.6 | 12.3 | 11.1 | 10.2 | 10.6 | 10.7 | 8.7 | 12.8 | 10.6 (1.2) |
| 45-49 | 18.8 | 16.6 | 19.6 | 18.9 | 17.2 | 17.2 | 19.7 | 17.7 | 17.5 | 17.3 | 14.8 | 14.1 | 17.4 (1.6) |
| 50-54 | 23.3 | 24.7 | 25.7 | 25.4 | 26.7 | 22.0 | 21.2 | 24.5 | 22.5 | 24.7 | 25.3 | 18.8 | 23.7 (2.2) |
| 55-59 | 33.4 | 29.0 | 32.2 | 31.0 | 29.0 | 29.6 | 28.9 | 27.8 | 32.8 | 30.0 | 29.7 | 29.5 | 30.3 (1.7) |
| 60-64 | 34.4 | 40.8 | 42.4 | 39.1 | 38.0 | 36.4 | 38.1 | 29.1 | 33.4 | 31.7 | 38.0 | 30.8 | 36.0 (4.0) |
| 65-69 | 45.4 | 48.0 | 53.2 | 47.8 | 48.0 | 44.9 | 43.3 | 46.4 | 47.6 | 47.2 | 38.7 | 39.7 | 45.8 (3.7) |
| 70-74 | 52.2 | 55.1 | 52.5 | 47.5 | 60.3 | 56.9 | 63.4 | 56.9 | 54.4 | 62.1 | 60.1 | 49.2 | 55.9 (4.8) |
| 75-79 | 60.8 | 72.6 | 70.7 | 69.0 | 55.6 | 68.9 | 61.3 | 66.3 | 74.3 | 69.1 | 71.6 | 83.2 | 68.6 (6.9) |
| 80-84 | 83.2 | 81.8 | 91.7 | 85.8 | 102.4 | 88.8 | 95.6 | 84.7 | 99.6 | 77.5 | 73.6 | 90.8 | 88.0 (8.3) |
| 85+ | 185.3 | 175.1 | 192.0 | 153.2 | 170.4 | 173.4 | 139.1 | 164.8 | 150.5 | 139.6 | 142.1 | 144.3 | 160.8 (17.7) |

